# Variation in the Prevalence of Postpartum Depressive Symptoms in Brazil: National Health Survey, 2013 and 2019

**DOI:** 10.1101/2025.07.14.25331529

**Authors:** Carolina Teixeira Silva, Claudia de Souza Lopes, Claudia Reis Miliauskas

## Abstract

Postpartum depression (PPD) is considered a public health issue due to its prevalence and its impact on the health of women and children [4,5]. A literature review of studies published between 2009 and 2022 on the Brazilian population regarding the prevalence and associated factors of PPD found that the prevalence of PPD ranged from 7.2% in Recife to 50.6% in Salvador [5]. The main objective of this study is to analyze the variation of postpartum depression in Brazil in the years 2013 and 2019. Additionally, the specific objectives are to calculate the prevalence of postpartum depression in Brazil and to evaluate the variation of postpartum depression according to socioeconomic, demographic, and regional characteristics. This study utilized data from two population-based, cross-sectional surveys: the National Health Survey (PNS) conducted in 2013 and 2019. The primary outcome assessed was postpartum depressive symptoms in women aged 18 years or older who had given birth within the 18 months prior to the interview date. The evaluation of depressive symptoms was based on the Patient Health Questionnaire-9 (PHQ-9), with a cutoff score of 10 or higher indicating the presence of significant depressive symptoms. The variation in prevalence between the 2013 and 2019 surveys was expressed both in absolute terms and as a percentage change of the prevalence ratio. Notably, there was a shift in the demographic profile of the women, particularly concerning age and educational attainment. An increase in the prevalence of postpartum depressive symptoms was observed across all variables and categories studied, with a relative increase of 41.3% in Brazil, rising from 9.9% in 2013 to 14% in 2019. When evaluating the selected age groups, it is noteworthy that the greatest relative variation in prevalence was among women aged 35 years or older (148.4%). Additionally, a high variation was found for the lowest level of education considered—up to incomplete primary education (94.7%). This study demonstrated an increase in postpartum depressive symptoms between 2013 and 2019 in Brazil. Among the main explanations for these increases are the economic crisis that occurred in the country during this period and changes in Brazilian family structures. Therefore, it is important to highlight the need for public investment in maternal mental health during the postpartum period.

## Introduction

According to the Pan American Health Organization (PAHO), depression is the leading cause of disability worldwide, affecting over 300 million people across all age groups. Data from the Global Burden of Disease Study indicate that depression is among the top three causes of years lived with disability, particularly among women (GBD, 2017).

In Brazil, a study assessing the trend in the prevalence of depressive symptoms in the general population between 2013 and 2019 revealed an increase from 7.9% to 10.8%. This increase, both in overall prevalence and in absolute and relative increments, was more pronounced among women (LOPES et al., 2022). Consistent with this, a systematic review and meta-analysis conducted in 2014 with Brazilian adults concluded that the prevalence of depression is up to twice as high in women compared to men (SILVA et al., 2014).

Postpartum depression (PPD) is a common disorder in women following childbirth and can present with a variety of symptoms and consequences. Furthermore, it is considered a public health issue due to its high prevalence and impact on the health of women and children. This disorder typically begins one month after the birth of the baby and can last for years postpartum (BOTTINO, 2011; WOOLHOUSE et al., 2014; THEME FILHA et al., 2016).

Three of the most common psychiatric disorders affecting women during the perinatal period, categorized by severity, are postpartum blues, postpartum depression, and postpartum psychosis. Postpartum blues occurs within the first two weeks after childbirth and is characterized as a transient mood depression, typically brief and relatively benign, with main symptoms including mood swings, tearfulness, and irritability. In contrast, postpartum psychosis generally manifests within the first three weeks postpartum and is marked by high severity and intensity of symptoms, which may include delusions and hallucinations, posing significant risks to both the mother and the newborn (MARTINS, 2006). Additionally, it is important to distinguish between perinatal depression, which occurs throughout the entire pregnancy and postpartum period, and postpartum depression (PPD), which specifically occurs after childbirth.

In Brazil, the most recent national data available come from the Nascer no Brasil survey conducted between 2011 and 2012, a hospital-based study that found a prevalence of postpartum depressive symptoms of 26.3% within the 6 to 18 months postpartum period (THEME FILHA et al., 2016). According to a literature review of studies published between 2009 and 2022 on the Brazilian population regarding the prevalence and associated factors of PPD, prevalence rates ranged from 7.2% in Recife to 50.6% in Salvador. The variability in these results can be attributed to differences in the timing of assessments, the various screening instruments used, the cutoff points applied, as well as disparities in access to prenatal and postnatal care, cultural factors, and socioeconomic differences among study populations (SANTANA et al., 2022).

Wang et al. (2021) conducted a systematic review and meta-analysis investigating the use of the Patient Health Questionnaire-9 (PHQ-9) for screening perinatal depression. Perinatal depression is a prevalent and disabling problem during pregnancy and the postpartum period. Among the main findings of the study are that the PHQ-9 has good sensitivity and specificity for screening perinatal depression and that its performance is comparable to the Edinburgh Postnatal Depression Scale (EPDS), which, until now, is the most recommended and widely used scale for perinatal depression. The study concludes that the PHQ-9 is a viable option for screening depression during pregnancy and postpartum periods.

In Brazil, so far, the national-level studies that have assessed women’s reproductive health include the National Demographic and Health Survey (PNDS), which has limitations for direct comparison across its different editions due to methodological changes and different institutions conducting each survey (Agência Notícias IBGE, 2023). Additionally, there is the Nascer no Brasil study, a hospital-based research (THEME FILHA et al., 2016), and the National Health Survey (PNS), a household survey that is the first national study focused exclusively on evaluating the health status of the Brazilian population. In both editions of the PNS (2013 and 2019), symptoms of depression were assessed, and among women aged 18 and older who had given birth within 18 months prior to the interview, the prevalence of postpartum depressive symptoms was investigated. This approach allowed for the assessment of depression prevalence in the postpartum period among Brazilian women.

To the best of our knowledge, this is the first population-based study with representative data for Brazil and its regions that analyzes the temporal variation of postpartum depressive symptoms. Therefore, the aim of this study is to assess how the prevalence of postpartum depressive symptoms varied in Brazil between 2013 and 2019, as well as to examine how this prevalence may have changed across different socioeconomic, demographic, and macro-regional groups within the country.

## Methods

### Study Design and Sample

This study utilized data from two editions of the National Health Survey (PNS), conducted in 2013 and 2019. These are cross-sectional, population-based household surveys. The surveys are nationally representative and were carried out by the Ministry of Health (MS) in partnership with the Brazilian Institute of Geography and Statistics (IBGE). The primary objective of the PNS is to provide data on the health status and lifestyle of the Brazilian population, including information on access to and utilization of health services, continuity of care, healthcare financing, and preventive actions.

The National Sample Survey of Households (PNS) was developed to provide representative estimates for the entire Brazilian territory, with the target population consisting of residents of permanent private households. The two editions of the PNS, conducted in 2013 and 2019, employed a three-stage conglomerate sampling design. As part of the Integrated System of Household Surveys (SIPD), the first stage of selection was based on the Master Sample. In the second stage, households were selected through simple random sampling within each primary sampling unit. Finally, in the third stage, also using simple random sampling, one resident was chosen (aged 18 or older in the 2013 edition and 15 or older in the 2019 edition) to respond to the specific questionnaire. Additionally, expansion factors were defined for the primary sampling units, households, all residents, and the selected individual, ensuring that the results are representative of the entire Brazilian population.

The first edition of the survey (PNS 2013) was conducted between August and November 2013, while the second edition (PNS 2019) took place in the field in August 2019, with data collection concluding in March 2020. According to IBGE, a total of 81,357 households were selected in 2013, and 108,525 in 2019. The calculated non-response rate for the common questionnaire administered to all residents was 20.8% in 2013 and 13.2% in 2019, remaining below the planned rates of 23% in 2013 and 20% in 2019. When considering the specific questionnaire for selected residents, non-response rates were slightly higher, at 25.9% in 2013 and 16.2% in 2019 (IBGE, 2015; IBGE, 2020).

### Variable description

For comparability between the two surveys, this study employed as an eligibility criterion only women aged 18 years or older who had given birth within the previous 18 months prior to the interview date.

The dictionary file and microdata were obtained via the IBGE’s official website. After analyzing the questionnaire, the variables of interest for this study were identified, followed by data processing.

The geographic variables included: Brazil and its macroregions (“North,” “Northeast,” “Midwest,” “Southeast,” and “South”) and household location (“urban” or “rural”). The sociodemographic variables considered were: age group (18 to 24 years, 25 to 34 years, and 35 years or older), race or color (white, black, or brown), educational level (no education and incomplete primary education, complete primary education and incomplete high school, complete high school or higher), marital status (“single, divorced, or widowed not cohabiting” and “married or cohabiting”). Additionally, parity was evaluated, defined as the number of previous births by the woman, categorized as ‘1 birth’ or ‘2 or more births’.

### Assessment of postpartum depressive symptoms

The assessment of postpartum depressive symptoms was conducted using the PHQ-9. This instrument consists of a questionnaire with nine items, each referring to a symptom associated with depression according to the diagnostic criteria established by the Diagnostic and Statistical Manual of Mental Disorders, Fourth Edition (DSM-IV). All questions offer four response options: “not at all,” “several days,” “more than half the days,” and “nearly every day,” corresponding to scores of 0, 1, 2, and 3 respectively. Consequently, the minimum possible score is 0 and the maximum is 27 (Lopes et al., 2016).

The cutoff point adopted in this study aligns with that used by Wang et al. (2021), who found comparable performance between the EPDS and PHQ-9; they considered a PHQ-9 score of 10 or higher as indicative of clinically significant depressive symptoms.

### Data Analysis

Prevalence estimates for the 2013 and 2019 National Health Survey (PNS) will be calculated along with their respective 95% confidence intervals. The complex sampling design of the survey and its population weights will be incorporated into all estimates. Changes in prevalence between the 2013 and 2019 surveys will be expressed as both absolute differences and percentage variations of the prevalence ratio. Both types of variation will be estimated using generalized linear models, employing a Gaussian model for the estimation of absolute differences and a Poisson model for the percentage change in prevalence ratio. Data manipulation will be performed using two statistical software packages: SAS and R, utilizing procedures “descriptives” and “survey,” respectively, which account for the effects of complex sampling in the analysis process.

### Ethical Aspects

This study will utilize secondary data from the 2013 and 2019 PNS, which were approved by the National Research Ethics Committee (CONEP) of the National Health Council (CNS), in June 2013 (Ordinance No. 328.159) and August 2019 (Ordinance No. 3.529.376), respectively. All participants were informed about the survey and provided written informed consent at the time of the interview.

## Results

The first table presents the sociodemographic and regional characteristics of women aged 18 years or older who had given birth within 18 months prior to the interview date, based on the samples from the National Health Survey (PNS) of 2013 and 2019. A total of 1,927 women were interviewed. Overall, there was little change in the profile of women who gave birth over the six-year period evaluated, except for age group and educational level. Regarding age, a significant variation was observed in the youngest group (18-24 years), with a reduction in parity prevalence from 37.3% to 28.6%, and an increase in the oldest group (35 years or more), which rose from 15.7% to 23.5%. Concerning educational level, a significant change was noted in parity among women with at least completed secondary education or higher, increasing from 52.8% to 61.6%.

Table 2 displays the prevalence of postpartum depressive symptoms among women aged 18 years or older who had given birth within 18 months prior to the survey, according to sociodemographic characteristics and previous number of births. Additionally, absolute and relative variations in these prevalences between 2013 and 2019 are presented. An increase in postpartum depressive symptoms prevalence was observed across all variables and categories studied, with a relative increase of 41.3% nationally, rising from 9.9% in 2013 to 14% in 2019.

**Table 1:** Women aged 18 years or older who had a child within 18 months prior to the interview date, categorized by sociodemographic characteristics and total number of births.

|  |  | 2013 |  |  | 2019 |  |  |
| --- | --- | --- | --- | --- | --- | --- | --- |
|  |  | n | N | % (CI95%) | n | N | % (CI95%) |
| Brazil | Total | 1927 | 3.925.807 | 100 | 2069 | 3.794.600 | 100 |
| Macroregions | North | 542 | 402.354 | 10,2(8,8;11,9) | 491 | 420.202 | 11,1(9,7;12,6) |
|  | Northeast | 581 | 1.117.469 | 28,5(25,4;31,7) | 775 | 1.126.950 | 29,7(26,9;32,7) |
|  | Southeast | 363 | 1.496.865 | 38,1(34,4;42) | 352 | 1.409.735 | 37,2(33,2;41,3) |
|  | South | 202 | 585.679 | 14,9(12,6;17,6) | 202 | 493.603 | 13(11,1;15,1) |
|  | Midwest | 239 | 323.440 | 8,2(7,1;9,5) | 249 | 344.110 | 9,1(7,6;10,8) |
| Household situation | Urban | 1544 | 3.303.620 | 84,2(81,5;86,5) | 1519 | 3.200.309 | 84,3(82,2;86,3) |
|  | Rural | 383 | 622.187 | 15,8(13,5;18,5) | 550 | 594.291 | 15,7(13,7;17,8) |
| Maternal age | 18-24 | 655 | 1.464.972 | 37,3(33,8;40,9) | 558 | 1.050.586 | 28,6(25,3;32,2) |
|  | 25-34 | 973 | 1.846.166 | 47(43,5;50,6) | 995 | 1.756.513 | 47,9(44,1;51,6) |
|  | 35 or more | 299 | 614.669 | 15,7(13,3;18,4) | 448 | 862.805 | 23,5(20,3;27) |
| Skin color or ethnicity <sup>a</sup> | White | 611 | 1.553.974 | 39,6(36;43,3) | 599 | 1.376.875 | 36,3(32,7;40) |
|  | Black or mixed-race | 1276 | 2.322.238 | 59,2(55,4;62,8) | 1423 | 2.357.735 | 62,1(58,4;65,7) |
| Level of education | No schooling or some elementary school | 479 | 912.002 | 23,2(20,5;26,2) | 471 | 718.653 | 18,9(16,5;21,7) |
|  | Elementary school or some high school | 466 | 941.786 | 24(20,9;27,3) | 444 | 737.927 | 19,4(16,9;22,3) |
|  | High school or more | 982 | 2.072.019 | 52,8(49;56,5) | 1154 | 2.338.020 | 61,6(58,1;65) |
| Marital Status | Non-cohabiting | 387 | 660.328 | 16,8(14,4;19,5) | 475 | 781.787 | 20,6(17,6;23,9) |
|  | Married or cohabiting | 1540 | 3.265.480 | 83,2(80,5;85,6) | 1594 | 3.012.812 | 79,4(76,1;82,4) |
| Parity | 1 | 715 | 1.549.350 | 39,5(35,9;43,1) | 718 | 1.502.676 | 39,6(36,3;43) |
|  | 2 or more | 1212 | 2.376.457 | 60,5(56,9;64,1) | 1351 | 2.291.924 | 60,4(57;63,7) |

95% Confidence Interval (CI): Lower Limit – Upper Limit. n: sample size. N: estimated number considering sampling weights. <sup>a</sup> Categories "Yellow" and "Indigenous" were excluded.

**Table 2:** Prevalence of postpartum depressive symptoms in women aged 18 or over who had a child up to 18 months before the date of the interview and according to sociodemographic characteristics, absolute prevalence difference and relative prevalence difference.

|  |  | 2013 | 2019 | Absolute prevalence difference | Relative prevalence difference |
| --- | --- | --- | --- | --- | --- |
|  |  | %(CI95%) | %(CI95%) | %(CI95%) | %(CI95%) |
| Brazil | Total | 9,9(7,8;12,5) | 14(11,7;16,7) | 4,1(0,6;7,5) | 41,3(5;90,2) |
| Macroregions | North | 12,5(7,8;19,5) | 13,7(9,3;19,8) | 1,2(-6,7;9,1) | 9,8(-40,2;101,4) |
|  | Northeast | 10,8(7,1;16,3) | 12,6(9,6;16,3) | 1,7(-3,9;7,3) | 15,7(-29,1;89) |
|  | Southeast | 7,1(4;12,2) | 14,7(10,2;20,8) | 7,6(1;14,2) | 107,6(7,3;301,5) |
|  | South | 11,2(6,8;18,1) | 14,1(9;21,4) | 2,9(-5,4;11,1) | 25,5(-35,5;144,2) |
|  | Midwest | 14(7,8;23,6) | 15,9(10,3;23,6) | 1,9(-9,1;12,9) | 13,7(-45,8;138,4) |
| Maternal age | 18-24 | 11,8(8,2;16,6) | 15,9(11,7;21,2) | 4,1(-2,2;10,5) | 34,9(-15,1;114,5) |
|  | 25-34 | 9,6(6,7;13,4) | 12,4(9,5;15,9) | 2,8(-1,8;7,4) | 29,1(-16,3;99,2) |
|  | 35 or more | 6,4(3,5;11,3) | 15,8(10,6;23) | 9,5(2;16,9) | 148,4(21,8;406,5) |
| Skin color or ethnicity <sup>a</sup> | White | 9,2(6,2;13,4) | 12,1(8,4;17,2) | 2,9(-2,8;8,7) | 32(-23,1;126,7) |
|  | Black or mixed-race | 10,5(7,8;13,9) | 15,1(12,3;18,4) | 4,6(0,4;8,8) | 44,2(2,2;103,3) |
| Level of education | No schooling or some elementary school | 8,8(5,9;13,2) | 17,2(11,7;24,7) | 8,4(0,9;15,9) | 94,7(11,3;240,4) |
|  | Elementary school or some high school | 11,8(7,3;18,6) | 16,1(11,9;21,3) | 4,2(-3,2;11,7) | 35,5(-22,6;137,1) |
|  | High school or more | 9,5(6,8;13,1) | 12,3(9,6;15,8) | 2,9(-1,5;7,3) | 30,3(-14;97,2) |
| Marital Status | Non-cohabiting | 14,1(9,5;20,4) | 20,2(14;28,3) | 6,1(-3;15,1) | 43,2(-15,5;142,7) |
|  | Married or cohabiting | 9(6,8;11,9) | 12,4(10,2;15) | 3,3(-0,2;6,9) | 36,8(-3,2;93,3) |
| Parity | 1 | 9,6(6,4;14,1) | 10(7,1;13,9) | 0,4(-4,6;5,4) | 4,5(-37,3;74,1) |
|  | 2 or more | 10,1(7,5;13,4) | 16,6(13,4;20,4) | 6,5(1,8;11,1) | 64,3(14,4;135,9) |

95% Confidence Interval (CI): Lower Limit – Upper Limit. <sup>a</sup> Categories "Yellow" and "Indigenous" were excluded.

Regarding the major regions of Brazil analyzed, the highest relative variation in prevalence was found in the Southeast region (107.6%), while the lowest was observed in the North region (9.8%). When examining specific age groups, it is noteworthy that the greatest relative variation occurred among women aged 35 years or older (148.4%). Furthermore, a substantial increase was identified among women with up to incomplete fundamental education—the lowest educational level considered—showing an increase of 94.7%.

When considering the skin color/race of women, no significant variation was observed across any of the evaluated categories (white, black, or mixed-race). Similarly, the results were consistent when analyzing the variable of marital status (not cohabiting, married, or cohabiting).

Regarding parity, there was a 64.3% increase in the prevalence of postpartum depressive symptoms among women with two or more births, rising from 10.1% in 2013 to 16.6% in 2019. In contrast, the increase among primiparous women was modest, with a variation of 4.5%, from 9.6% in 2013 to 10% in 2019.

It is important to note that due to the sample size limitations for the rural category within the area of residence variable, it was not possible to perform this analysis for calculating prevalences of depressive symptoms and their variations in that subgroup.

## Discussion

This study is the first to compare the overall landscape of postpartum depressive symptoms in Brazil across two distinct periods. The estimates derived from the PHQ-9 indicated a relative increase of 41.3% in the prevalence of postpartum depressive symptoms between 2013 and 2019, rising from 9.9% to 14%, respectively. Furthermore, this increase was more pronounced among women residing in the Southeast region of the country, those aged 35 years or older, women with lower educational attainment (up to incomplete fundamental education), and multiparous women (with two or more births).

Regarding the profile of women who gave birth in Brazil between 2013 and 2019, the findings align with both global and national trends, which show that women are having children later (35 years or older) and with higher levels of education. The primary factor associated with this shift appears to be the increasing participation of women in the labor market (IBGE, 2024; Budds et al., 2016; Martin et al., 2019). Additionally, there was a significant reduction in the proportion of women giving birth between ages 18 and 24 from 2013 to 2019, suggesting improvements in access to information and sexual and reproductive health services. This progress is directly related to expanded coverage of Primary Health Care and the expansion of the Family Health Strategy, which bring young adults closer to healthcare professionals (IBGE, 2024; Giovanella et al., 2021; MS, 2020).

In 2022, a trend study on depression in Brazil was conducted, which found an increase in depression prevalence from 39.8% between 2013 and 2019 among women aged 18 years or older (Lopes et al., 2022). This increase is similar to the 41.3% rise in postpartum depression prevalence identified in the present study. These findings align with a trend study of depression and postpartum depression conducted in Denmark between 2000 and 2022 (Egsgaard et al., 2024), which concluded that the prevalence of depression and postpartum depression follow similar temporal trends.

A systematic review on the most common determinants affecting perinatal mental health in low- and middle-income countries observed that economic risk is broadly associated with an increased risk of common perinatal mental disorders (Fisher et al., 2012). The economic crisis initiated in Brazil in 2014, the year between the two editions of the PNS, affected all regions of the country, with a greater impact on the labor market in the Southeast region (Mattei, 2020). Generally, periods of crisis are felt more intensely by vulnerable groups, especially in Brazil where no effective social protection measures were implemented during this period. Therefore, the economic crisis experienced in Brazil during this time may partially explain the increase in postpartum depression prevalence among women with lower educational levels (94.7%) and the significant rise observed in the Southeast region of the country (107.6%).

Among the main findings of this study, the significant increase in postpartum depression prevalence among women aged 35 years or older stands out, with a rise of 148.4%. In contrast, the relative increase for other age groups evaluated was around 30%. According to the 2010 and 2022 demographic censuses, there was a change in the composition of Brazilian households during this period; previously, 38.7% of women were responsible for household management, and by 2022, this percentage increased to 49.1%, particularly among women aged 40 to 59 years. This shift led to an increased responsibility for these women (Census, 2022). Linked to the economic crisis affecting the period of this study and the rising responsibilities of women as household heads, there has also been an increase in what is called the “sandwich generation” among women both in Brazil and globally. The “sandwich generation” refers to women who are simultaneously caring for their children and aging parents (De Jesus & Wajnman, 2016). This phenomenon is related to increased life expectancy and delayed motherhood and predominantly affects women aged 35 to 49 years (Feijó, 2024). These aforementioned changes likely had a negative impact on the mental health of women aged 35 or older who recently became mothers, which may explain why this group experienced a greater increase in postpartum depression prevalence compared to other age groups.

The “Nascer no Brasil” study conducted in 2011/2012 found a prevalence of 26.3% of postpartum depressive symptoms in Brazil (Theme Filha et al., 2016), a result significantly higher than the 9.9% identified in the present study for 2013. One of the main reasons for this divergence is the use of different screening instruments for postpartum depressive symptoms. “Nascer no Brasil” employed the Edinburgh Postnatal Depression Scale (EPDS), which considers two anxiety symptoms that, although common in perinatal depression, are not part of the diagnostic criteria for depressive disorders. Conversely, the EPDS does not include four somatic symptoms associated with major depression—sleep disturbances, fatigue, weight or appetite changes, and agitation—that are considered in the PHQ-9 (Wang et al., 2021; Zhong et al., 2014). This is often cited as an advantage of the EPDS, as some of these somatic symptoms can be characteristic of postpartum states rather than indicative of depression itself. Additionally, the mode of interview differed between the two studies: in “Nascer no Brasil,” the EPDS was administered via telephone call with a response rate of 49.9%, whereas in the PNS 2013, the PHQ-9 was conducted through face-to-face interviews at participants’ homes, with a response rate of 74.1% for the specific questionnaire for selected residents. Furthermore, because the PNS interviews were conducted by interviewers face-to-face, responses may be subject to social desirability bias—the tendency for individuals to provide answers they perceive as socially acceptable rather than truthful or accurate—especially in regions where mental health remains stigmatized or misunderstood (Wang et al., 2021; Zhong et al., 2014; CRP, 2021). The fact that the interviewers in the PNS were of both sexes, unlike in the “Nascer no Brasil” study where interviewers were exclusively female, may also contribute to social desirability bias in responses. Besides the differences in the instruments used in each study, it is important to highlight the timing of data collection. The present study analyzed women between 0 and 18 months postpartum, whereas “Nascer no Brasil” considered women between 6 and 18 months postpartum.

Other studies comparing the EPDS and PHQ-9 have also found differences in screening for postpartum depressive symptoms when using these instruments. Larsen et al. (2024) compared the prevalence of postpartum depressive symptoms among women in Kenya six weeks after childbirth, using four different instruments—including both the EPDS and PHQ-9—and found a higher prevalence with the EPDS at 9.2%, compared to only 3.2% with the PHQ-9. Notably, the prevalence identified by the EPDS was nearly three times higher than that detected by the PHQ-9.

Another study comparing the use of EPDS and PHQ-9 against DSM-IV diagnoses concluded that the EPDS has better accuracy than the PHQ-9 for identifying women with postpartum depression symptoms (Hanusa et al., 2008). These findings underscore that different screening tools and methodological approaches can significantly influence prevalence estimates of postpartum depression, highlighting the importance of instrument selection and timing in epidemiological studies.

Liu et al. (2024), in their comparison of the Edinburgh Postnatal Depression Scale (EPDS) and the Patient Health Questionnaire-9 (PHQ-9) among pregnant women, found that psychological symptoms may be more significant in the assessment of depressive symptoms when using the EPDS, whereas physical symptoms tend to have a greater influence in the PHQ-9. Despite these differences, other studies have concluded that both instruments can be effectively utilized for screening postpartum depression symptoms (Wang et al., 2021; Srisurapanont et al., 2023).

When comparing the results of the present study with international literature that employed the PHQ-9 for postpartum depression screening, a study published in 2014 conducted in Mexico City, Mexico, involving 210 women, reported a prevalence of postpartum depression (PPD) of 20.0% within six months postpartum (Ma. Asunción Lara et al., 2014). This prevalence is notably higher than the 9.9% observed in this study for 2013. Conversely, another investigation carried out in Minnesota, United States, in 2016 with a sample of 6,523 women found a postpartum depression prevalence of 10.7% within three months after childbirth (Sidebottom et al., 2023), a result consistent with the findings of this study. Furthermore, a systematic review of articles published between 2005 and 2014 identified higher prevalence rates in developing countries compared to developed nations. However, this review did not find any studies from developing countries utilizing the PHQ-9; only six studies from developed countries employing this instrument reported postpartum depressive symptom prevalence rates ranging from 3.4% to 12.5% within six months postpartum (Norhayati et al., 2015). These comparisons highlight how factors such as socioeconomic context, timing of assessment, and choice of measurement instrument can significantly influence estimates of postpartum depression prevalence across different populations and international studies.

## Strengths and Limitations

This study is the first to assess the variation of postpartum depression in Brazil using data from two editions of a nationally representative survey. This approach enabled the comparison of variations according to population characteristics.

Among the limitations of this study is the sample size, which prevented the evaluation of the “area of residence” variable (urban versus rural). Additionally, there were changes in the collection of income and occupation data between the two editions of the National Health Survey (PNS), which hindered the assessment of these variables. Including such variables would have enriched the analysis and contributed to a more in-depth understanding of the findings.

## Conclusions

The present study demonstrated an increase in postpartum depressive symptoms in Brazil between 2013 and 2019. Notably, there was a significant rise among women over 35 years of age. Additionally, considerable variation was observed in the Southeast region of the country and among women with lower educational levels. Among the primary explanations for these increases are the economic crisis that occurred in Brazil during the period between the survey editions and changes in Brazilian family structures. Therefore, it is essential to emphasize the need for public investment in maternal mental health during the postpartum period.

## Data Availability

All data produced are available online at IBGE's official website.

## Notes

### Competing Interest Statement

The authors have declared no competing interest.

### Funding Statement

This study did not receive any funding

### Author Declarations

This study will utilize secondary data from the 2013 and 2019 PNS, which were approved by the National Research Ethics Committee (CONEP) of the National Health Council (CNS), in June 2013 (Ordinance No. 328.159) and August 2019 (Ordinance No. 3.529.376), respectively.

## Bibliographic References

OPAS. Depressão - OPAS/OMS | Organização Pan-Americana da Saúde. Disponível em: <https://www.paho.org/pt/topicos/depressao>.

GBD 2017 Disease and Injury Incidence and Prevalence Collaborators. Global, regional, and national incidence, prevalence, and years lived with disability for 354 diseases and in­juries for 195 countries and territories, 1990-2017: a systematic analysis for the Global Burden of Disease Study 2017. Lancet 2018; 392:1789–858.

Lopes, C. De S. et al. Trend in the prevalence of depressive symptoms in Brazil: results from the Brazilian National Health Survey 2013 and 2019. Cadernos de Saúde Pública, v. 38, n. suppl 1, 2022.

Silva, M. T. et al. Prevalence of depression morbidity among Brazilian adults: a systematic review and meta-analysis. Revista Brasileira de Psiquiatria, v. 36, n. 3, p. 262–270, set. 2014.

Bottino, M, 2011. Universidade do Estado do Rio de Janeiro. Disponível em: <https://www.bdtd.uerj.br:8443/bitstream/1/4056/1/T1287%20Marcella%20Bottino%20completa%20protegida.pdf>. Acesso em: 22 abr. 2025.

Woolhouse, H. et al. Maternal depression from early pregnancy to 4 years postpartum in a prospective pregnancy cohort study: implications for primary health care. BJOG: An International Journal of Obstetrics & Gynaecology, v. 122, n. 3, p. 312–321, 21 maio 2014.

Themme Filha, M. M., Ayers, S., Gama, S. G. N. D. and Leal, M. D. C. (2016). Factors associated with postpartum depressive symptomatology in Brazil: The Birth in Brazil National Research Study, 2011/2012. Journal of Afective Disorders, 194, pp. 159–167. doi: 10.1016/j.jad.2016.01.020

Martins Ra. A depressão materna do pós-parto: algumas compreensões e outros nevoeiros [dissertação]. São Paulo: Pontifícia Universidade Católica de São Paulo; 2006

Santana, G. W. et al. Prevalência e fatores de risco da depressão pós-parto no Brasil: uma revisão integrativa da literatura. Debates em Psiquiatria, v. 12, p. 1–23, 3 nov. 2022.

Wang, L. et al. Screening for perinatal depression with the Patient Health Questionnaire depression scale (PHQ-9): A systematic review and meta-analysis. General Hospital Psychiatry, v. 68, n. 1, p. 74–82, jan. 2021.

Agência de Notícias, 2023. PNDS vai a campo coletar informações sobre demografia, saúde reprodutiva e nutrição das crianças Agênciap de Notícias. Disponível em: <https://agenciadenoticias.ibge.gov.br/agencia-noticias/2012-agencia-de-noticias/noticias/38058-pnds-vai-a-campo-coletar-informacoes-sobre-demografia-saude-reprodutiva-e-nutricao-das-criancas>.

Instituto Brasileiro de Geografia e Estatística (IBGE). Pesquisa Nacional de Saúde 2013: Acesso e utilização dos serviços de saúde, acidentes e violências. Rio de Janeiro: IBGE; 2015. Brasil, Grandes Regiões e Unidades da Federação

IBGE. Pesquisa Nacional de Saúde 2019. Ciclos de Vida. Rio de Janeiro, 2021. Disponível em: https://biblioteca.ibge.gov.br/index.php/biblioteca-catalogo?view=detalhes&id=2101846.

IBGE. Pesquisa Nacional de Saúde. 2019. Conceitos e Métodos. Instrumentos de Coleta. PNS – Manual de Entrevista de Saúde. Disponível em: https://www.ibge.gov.br/estatisticas/sociais/saude/9160-pesquisa-nacional-desaude.html?=&t=conceitos-e-metodos. Acesso em 03 de Jun. de 2025.

Lopes, C. S. et al. Inequities in access to depression treatment : results of the Brazilian National Health Survey – PNS. International Journal for Equity in Health, v. 15, n. 154, p. 1–8, 2016.

IBGE, 2024. Estatísticas de Gênero Estudos e Pesquisas, Informação Demográfica e Socioeconômica, n.38. Disponível em: https://biblioteca.ibge.gov.br/visualizacao/livros/liv102066_informativo.pdf

Budds K, Locke A and Burr V (2016) “For some people it isn’t a choice, it’s just how it happens”: Accounts of ‘delayed’ motherhood among middle-class women in the UK. Feminism and Psychology. 26(2): 170–187.

Martin, J. A. et al. Births: Final Data for 2019. National Vital Statistics Reports: From the Centers for Disease Control and Prevention, National Center for Health Statistics, National Vital Statistics System, v. 70, n. 2, p. 1–51, 1 abr. 2021.

Giovanella, L. et al. Cobertura da Estratégia Saúde da Família no Brasil: o que nos mostram as Pesquisas Nacionais de Saúde 2013 e 2019. Ciência & Saúde Coletiva, v. 26, n. suppl 1, p. 2543–2556, jun. 2021.

Ministério da Saúde, 2020. Principais ações em saúde para prevenção da gravidez na adolescência. Disponível em: & https://www.gov.br/saude/pt-br/composicao/saps/noticias/2020/fevereiro/principais-acoes-em-saude-para-prevencao-da-gravidez-na-adolescencia>.

Egsgaard S, Bliddal M, Rasmussen L, Mægbæk ML, Liu X, Munk-Olsen T. Time trends in incidence of postpartum depression and depression in women of reproductive age. J Afect Disord. 2024 Dec 1;366:91–97. doi: 10.1016/j.jad.2024.08.167. Epub 2024 Aug 24. PMID: 39187186.

Fisher, J. et al. Prevalence and Determinants of Common Perinatal Mental Disorders in Women in low- and lower-middle-income countries: a Systematic Review. Bulletin of the World Health Organization, v. 90, n. 2, p. 139–149H, 24 nov. 2011.

Mattei T F, Cunha M S, 2020. A CRISE ECONÔMICA BRASILEIRA E SEUS EFEITOS SOBRE O EMPREGO FORMAL: UMA DECOMPOSIÇÃO SHIFT-SHARE ESTOCÁSTICA. Disponível em: <https://revistas.unila.edu.br/orbis/article/view/1781/1850>. Acesso em: 5 maio. 2025.

CENSO 2022: Em 12 anos, proporção de mulheres responsáveis por domicílios avança e se equipara à de homens | Agência de Notícias. Disponível em: <https://agenciadenoticias.ibge.gov.br/agencia-noticias/2012-agencia-de-noticias/noticias/41663-censo-2022-em-12-anos-proporcao-de-mulheres-responsaveis-por-domicilios-avanca-e-se-equipara-a-de-homens>.

De Jesus, J. C., & Wajnman, S. (2016). Geração sanduíche no Brasil: realidade ou mito? Revista Latinoamericana De Población, 10(18), 43–61.

Feijó, J, 2024. Proporção de mulheres da “Geração Sanduíche” fora do mercado é quase seis vezes maior do que a dos homens | Blog do IBRE. Disponível em: <https://blogdoibre.fgv.br/posts/proporcao-de-mulheres-da-geracao-sanduiche-fora-do-mercado-e-quase-seis-vezes-maior-do-que-dos?utm_source=portal-fgv&utm_medium=fgvnoticias&utm_id=fgvnoticias-2024-03-19>. Acesso em: 9 maio. 2025.

Zhong, Q. et al. Comparative performance of Patient Health Questionnaire-9 and Edinburgh Postnatal Depression Scale for screening antepartum depression. Journal of Afective Disorders, v. 162, p. 1–7, jun. 2014.

CRP, 2021. Janeiro Branco: falar sobre a saúde mental ainda é um tabu? - CRP19. Disponível em: <https://crp19.org.br/janeiro-branco-falar-sobre-a-saude-mental-ainda-e-um-tabu/>. Acesso em: 12 jun. 2025.

Ma. Asunción Lara, Laura Navarrete, Lourdes Nieto, Juan Pablo Barba Martín, José Luis Navarro, Héctor Lara-Tapia, Prevalence and Incidence of Perinatal Depression and Depressive Symptoms among Mexican Women, Journal of Afective Disorders, 10.1016/j.jad.2014.12.035.

Liu, W. et al. Comparison of the EPDS and PHQ-9 in the assessment of depression among pregnant women: Similarities and diferences. Journal of afective disorders (Print), v. 351, 1 jan. 2024.

Larsen A, Pintye J, Odhiambo B, Mwongeli N, Marwa MM, Watoyi S, Kinuthia J, Abuna F, Gomez L, Dettinger J, Bhat A, John-Stewart G. Comparing depression screening tools (CESD-10, EPDS, PHQ-9, and PHQ-2) for diagnostic performance and epidemiologic associations among postpartum Kenyan women: Implications for research and practice. J Afect Disord. 2023 Mar 1;324:637–644. doi: 10.1016/j.jad.2022.12.101. Epub 2022 Dec 29. PMID: 36586607; PMCID: PMC9990497.

Srisurapanont, M.; Oon-arom, A.; Suradom, C.; Luewan, S.; Kawilapat, S. Convergent Validity of the Edinburgh Postnatal Depression Scale and the Patient Health Questionnaire (PHQ-9) in Pregnant and Postpartum Women: Their Construct Correlations with Functional Disability. Healthcare 2023, 11, 699. 10.3390/healthcare11050699

Sidebottom AC, Vacquier M, LaRusso E, Schulte AK, Nickel A. Prenatal and postpartum depression diagnosis in a large health system: prevalence and disparities. Ann Med. 2023;55(2):2281507. doi: 10.1080/07853890.2023.2281507. Epub 2023 Nov 14. PMID: 37963220; PMCID: PMC10836261.

Norhayati, M. N. et al. Magnitude and risk factors for postpartum symptoms: A literature review. Journal of Afective Disorders, v. 175, p. 34–52, abr. 2015.

